# Clustering of >145,000 Symptom Logs Reveals Distinct Pre, Peri, and Post Menopausal Phenotypes

**DOI:** 10.1101/2023.12.12.23299821

**Authors:** Shravan G. Aras, Azure D. Grant, John P. Konhilas

## Abstract

**Background and Objectives:** The transition to menopause is commonly associated with disappearance of menstrual cycle symptoms and emergence of vasomotor symptoms. Although menopausal women report a variety of mood, digestive, and pain symptoms, it remains unclear what symptoms emerge prior to menopause, if symptoms occur in predictable clusters, how these clusters change from across the premenopause-perimenopause-menopause transition, or if distinct phenotypes are present within each life stage.

**Methods:** We present analysis of self-reported symptom presentation in premenopausal to menopausal women using the MenoLife app, which includes 4,789 (n=1,115(23%) premenopausal, n=1,388(29%) perimenopausal, n=2,286(48%) menopausal) individuals and 147,501 (n=27,371(19%) premenopausal, n=57,964(39%) perimenopausal, n=61,806(42%) menopausal) symptom logs. Clusters generated from logs of 45 different symptoms were assessed for similarities across methods: hierarchical clustering analysis (HCA), K-Means clustering of principal components of symptom reporting, and binomial network analysis. Participants were further evaluated based on menstrual cycle regularity or type of menopause.

**Results:** Menstrual cycle-associated symptoms (e.g., cramps, breast swelling), digestive, mood, and integumentary symptoms were characteristic of premenopausal women. Vasomotor symptoms, pain, mood, and cognitive symptoms were characteristic of menopause. Perimenopausal women exhibited both menstrual cycle-associated and vasomotor symptoms. Subpopulations across life stages presented with additional correlated mood and cognitive symptoms, integumentary complaints, digestive, nervous, or sexual symptoms. Symptoms also differed among women depending on the reported regularity of their menstrual cycles or the way in which they entered menopause. Notably, we identified a set of symptoms that were very common across life stages: fatigue, headache, anxiety, and brain fog. We consistently identified the lack of predictive power of hot flashes for other symptoms except night sweats.

****Conclusions**:** Together, premenopausal women exhibit menstrual cycle-associated symptoms and menopausal women reported vasomotor symptoms, perimenopausal women report both; and all report high rates of fatigue, headache, anxiety, and brain fog. Limiting focus of menopausal treatment to vasomotor symptoms, or to premenstrual syndrome in premenopausal women, neglects a large proportion of overall symptom burden. Future research and interventions targeting mood and cognitive, digestive, and integumentary symptoms are needed across stages of female reproductive life.

## Introduction

The symptoms attributed to any life stage must be viewed against an ever-evolving background of overall health, and those associated with the menstrual cycle and menopause may need an update. Menopausal symptoms have received increasing medical attention in the mid-19th century^1^, and premenstrual syndrome (PMS) has been medically acknowledged since at least 1935^2^. Although the baseline for female health (e.g., rate of obesity), and rate of reproductive health problems (e.g., polycystic ovarian syndrome) increased greatly from these eras, the clinical description of cycle-associated to climacteric symptoms has changed minimally. Health in America has degraded rapidly in the 21^st^ century, with well-documented increases in overweight and obesity^3,4^, metabolic disease^5,6^, and potentially poorer coping with perceived stress^7^, all of which can impact reproductive function^8,9^ and menopause^10–13^. In particular, the recent years of the COVID-19 pandemic comprise a uniquely stressful and isolating time, the health effects of which are still being desribed^14,15^. This changing background of general “health” in the population suggests that the experience of female reproductive life stages may differ from the reports in the 1990’s or even early 2000’s. Because life expectancy of women has now reached 80 (almost 35 years longer than at the turn of the 20th century^16^), a greater proportion (40%) of a woman’s lifespan is spent in menopause and, consequently, with menopausal symptoms. The interaction of the changing baseline health environment with aging physiology in a population in which menopausal women will become an estimated half of females by 2030^17^ creates a strong need for continuous study of symptoms and symptom clusters. These clusters can provide an “update” for clinicians as well as context for evaluating efficacy of new treatments for women of all ages.

Today, methods like symptom clustering have yielded objective diagnostic criteria and more specialized treatment in fields ranging from psychiatry^18^ to cardiology^19,20^, gastroenterology^21^, and female reproductive health^10,11,22–26^. Numerous efforts have been made to cluster symptoms among premenopausal to menopausal women using longitudinal clinical datasets and surveys^10,11,24,27–29^, and a few studies have employed self-collected data from menstrual tracking apps to evaluate premenopausal women^30,31^. Despite these efforts, lack of consensus remains regarding as to what: 1) symptoms modern-day women experience, 2) symptoms are consistently predictive of other symptoms, 3) symptom patterns are specific to each hormonal life stage, and 4) distinct phenotypes within life stages exist that may be served by different treatments.

Large-scale efforts have aimed to describe potential sub-groups of menopausal experience. Analytical attempts are frequently directed at the open-access *Study of Women’s Health Across the Nation* (SWAN) dataset^32^, which followed 3,289 patients at biannual office visits across 16 years, therefore capturing the full menopausal transition of many women but with low temporal resolution and heavy reliance on recall at the time of doctors’ appointments. These datasets have yielded many insights into the variety of symptoms experienced in menopause, and their potential trajectories and influential factors. However, they are yet to generate a consistent picture of “the menopausal experience” or even consistent “menopausal subtypes”. For example, Harlow et al., 2017^10^ used latent transition analysis to evaluate symptom relatedness in the SWAN dataset, identifying symptom severity clusters ranging from relatively asymptomatic to highly symptomatic. They additionally reported two distinct symptom cluster types: fatigue and psychosocial; VMS, sleep, and fatigue. A more recent analysis of a 557-woman subgroup with metabolic syndrom^11^, used latent class growth analysis to identify very different symptom clusters: sleep and urinary problems; VMS and vaginal dryness; and psychological, joint, and sexual dysfunction.

A separate menopausal cohort study^24^, the Women’s Wellness Research Study, identified a further set of symptom clusters using a single timepoint survey, psychological, fatigue, and sleep; VMS; pain and numbness; and panic attacks and racing heart. Moreover, the authors identified more and more severe symptoms in women with a history of breast cancer, who reported nearly double the rate of VMS and low libido. Woods et al., 2014^29^ drew on the Seattle Midlife Women’s Health Study, which collected annual surveys of 508 menopausal and perimenopausal women, identified only mildly symptomatic, moderately symptomatic, and highly symptomatic clusters. Finally, studies of populations around the world have suggested that culture and genetic background impact menopausal experience. An early study conducted via phone interview of 1,900 Chinese women identified lower prevalence of VMS, and a peak of symptoms during perimenopause^27^. Five symptom clusters were identified: muscular and GI pain; psychological, respiratory, VMS and sleep disturbance, and non-specific somatic (fatigue, dizziness, headache). Subsequent studies have confirmed lower incidence of VMS and higher pain reporting in Asian populations^33^. Additional differences may be present, with African American women exhibiting higher rates of VMS^34,35^, Caucasian and Asian women potentially experiencing higher rates of psychological symptoms^28^.

Considering the above-named studies, totaling over 7,000 women collectively, it remains unclear if consistent subtypes of menopause exist, and what physiological factors would underlie such cohort subtypes. Woods et al., 2014^29^, Harlow et al., 2017^10^, and Min et al., 2023^11^ all make clear that external health and socioeconomic factors (breast cancer history, financial stress, Caucasian or Asian ethnicity, and obesity/metabolic syndrome) can worsen the number and severity of symptoms, but do not provide consensus otherwise. Such different findings may be amplified by differences in data collection, analytical methods, number and range of reportable symptoms, symptom severity metrics, and changing perception of symptoms collected from different cultures in different decades.

In an attempt to reconcile results across analysis methods and life stages, we analyzed the characteristics of user-reported symptoms collected from a smartphone application designed to capture up to 45 symptoms in daily life. Analyzing symptom reporting across the continuum of *pre*- to *peri*- to *post*- menopausal enabled us to distinguish among symptoms dependent or independent of life stage. In addition, we aimed to avoid bias inherent in each of analytical method by employing several standard clustering methods: hierarchical agglomerative clustering (HCA) of symptom covariance, K-Means clustering of principal components generated from symptoms, and network analysis.

Drawing on existing reports, we hypothesized that most common symptoms among premenopausal women would be associated with the menstrual cycle (e.g., cramps, ovulation pain, breast swelling, spotting). We hypothesized that VMS would emerge in perimenopause, and menstrual-associated symptoms would disappear by menopause. Finally, we hypothesized that symptom patterning would vary by cycle regularity and by type of menopause. Comparison of premenopausal through postmenopausal populations enabled us to distinguish among symptoms that depend on life stage, and symptoms common to women independent of life stage. To address these hypotheses, as well as avoid bias inherent to each analytical method we looked for commonalities among the results of standard clustering methods: hierarchical agglomerative clustering (HCA) of symptom covariance, K-Means clustering of principal components generated from symptoms, and network analysis.

## Results

### Study Population

Using a smartphone-based application where participants choose from 45 climacteric conditions/symptoms, 25,369 users recorded a total of 447,802 symptoms. All symptoms were collected from Fall 2021 through Spring 2023 (**Supplemental Figure 1**). In addition to self-reported symptoms, participants also self-reported menopausal status using a series of onboarding questions in order to determine how menopause was entered and menstrual cycle regularity (if applicable).Using inclusion criteria outlined in **Figure 1**, a total of 4,789 out of the 25,369 total users and 147,501 symptoms out of the 447,802 total symptoms were included in the analysis. Of the 4,789 total women included in th analysis, 1,115 (23%) women met the criteria for premenopause, reporting a total of 27,731 symptoms (Table 1) with a median of 17 symptoms and median absolute deviation (MAD) of reported per user; 1,388 (29%) women met the criteria for perimenopause, reporting a total of 57,964 symptoms an increased and more variable median symptom rate of 23 (); 2,286 (48%) women met the criteria for menopause, reporting a total of 61,806 symptoms. Despite the increased number of menopausal users and symptoms, symptoms per user were remarkably consistent, with a median of 19 symptoms (). Note that some users did not answer if or how they had entered menopause (n=124) or logged chemotherapy (n=6). These users were excluded from the analysis (See **Figure 1**, **Table 1**) Distribution of symptom counts did not vary by group (**Figure 2**).

**Figure 1.**
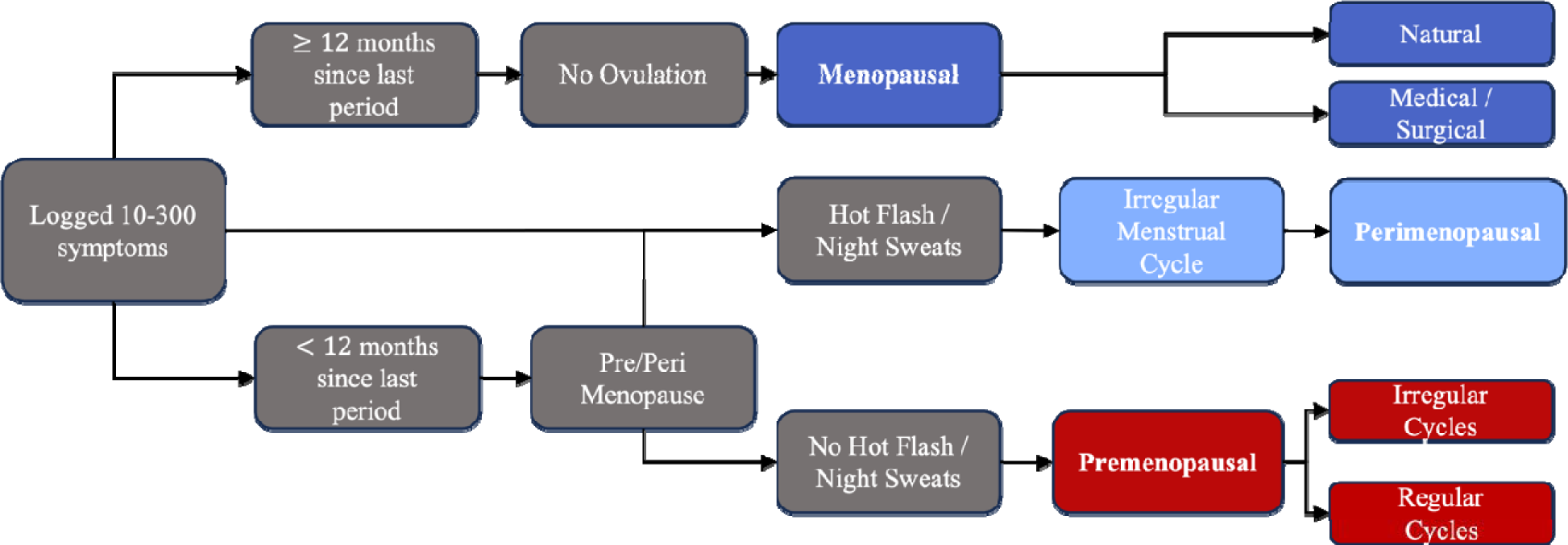
Inclusion criteria flow chart including separation of premenopausal, perimenopausal, and menopausal groups based on on-boarding survey questions asked in the MenoLife application.

**Figure 2.**
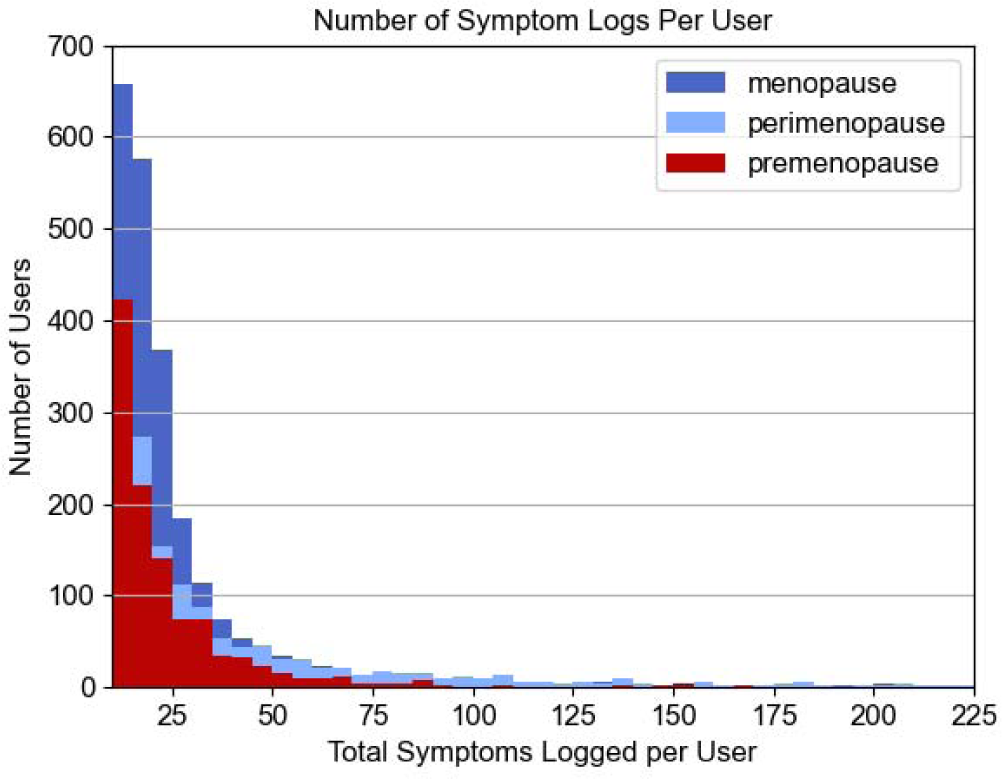
The number of users reporting each symptom count in premenopausal (red), perimenopausal (purple) and menopausal (blue) groups.

**Table 1.**
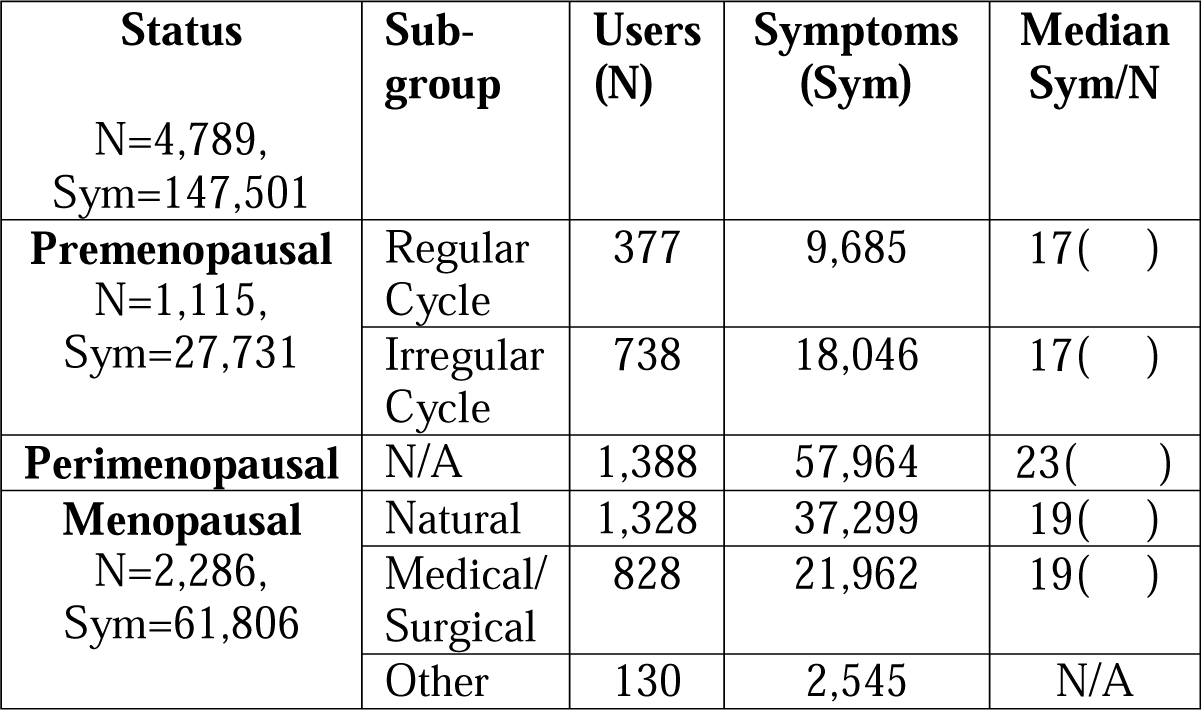
Number of women (N) comprising premenopausal, perimenopausal, and menopausal groups and their respective symptom counts (Sym). Sub-group others for menopausal women represents chemotherapy or unreported reason for menopause. These were excluded from analysis.

### Symptom Prevalence

The most common logs, presented as a percent of total logs, in premenopausal users were fatigue (7.94%), spotting (7.44%), cramps (6.55%), bloating (6.16%) and headaches (5.78%). By contrast, menopausal hot flashes greatly outweighed the prevalence of any other log (22.3%), followed by fatigue (5.13%), night sweats (4.31%), anxiety (3.96%), joint pain (3.52%), and bloating (3.45%). Perimenopausal women exhibited a combination of these most prevalent symptoms from the pre- and post-menopausal cohort with log prevalence of the following symptoms: hot flashes (14.8%), fatigue (6.33%), headaches and night sweats (each 4.77%), and cramps (4.38%) (**Figure 3A**).

**Figure 3.**
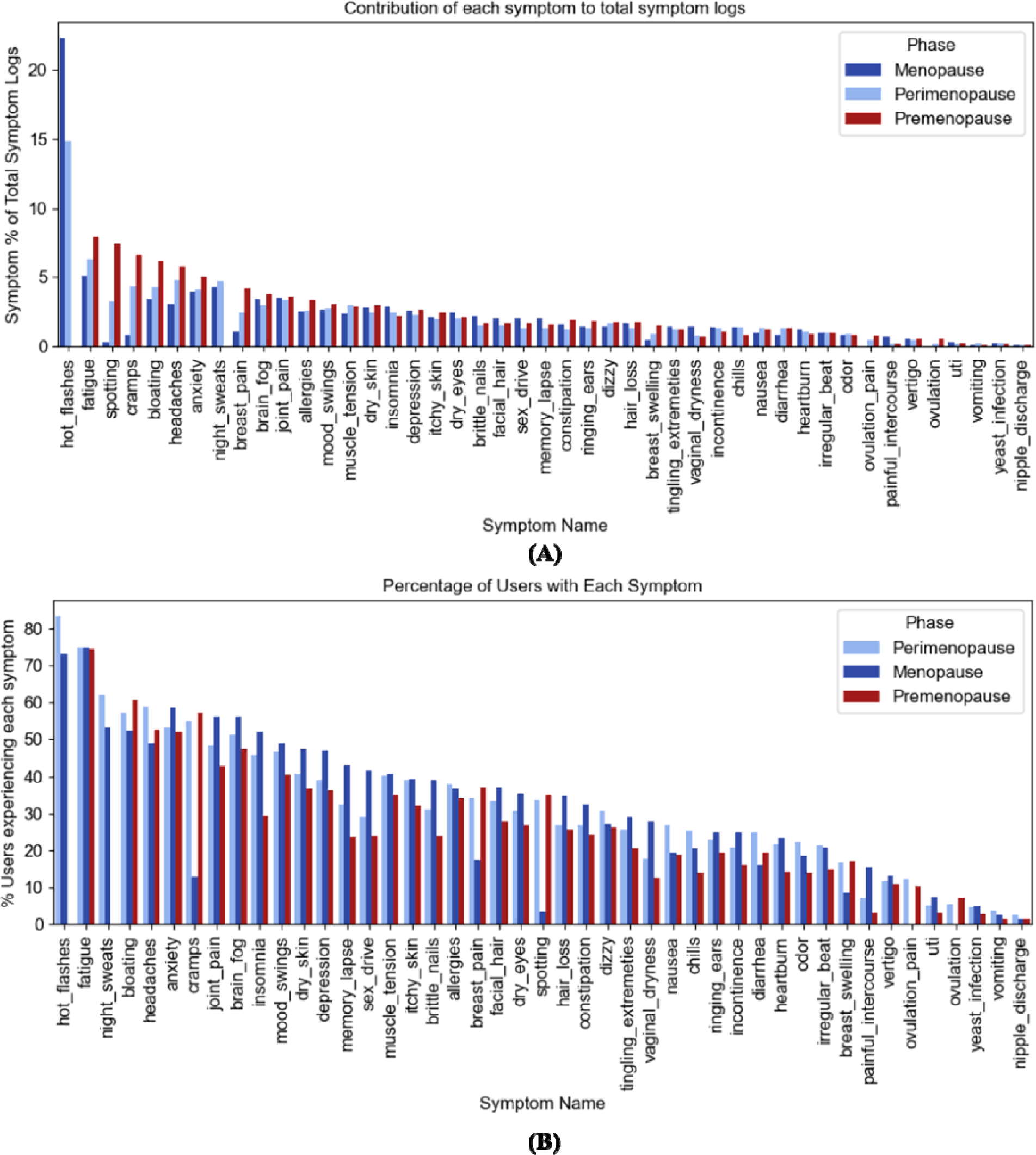
Symptom Reporting. (A) Histogram of each symptom as a percentage of total symptoms logged. **(B):** Histogram of the number of participant users reporting each symptom as a percentage of total participant users. Menstrual cycle-associated symptoms comprise most premenopausal logs (**red bars**) while VMS, pain, and brain fog distinguish menopausal (**purple**) users and perimenopausal users exhibit both menstrual cycle-associated and VMS **(blue bars)**.

The percentage of users reporting symptoms depended on self-reported life stage. Premenopausal women were most likely to report fatigue (74.4%), followed by bloating (60.6%), cramps (57.3%), headaches (52.6%), and anxiety (52.2%), which closely mimicked the most common logs.

Perimenopausal women exhibited the highest rate of hot flashes (83.4%) and night sweats (62.2%), followed by fatigue (74.8%), headaches (58.9%), and bloating (57.1%), all comparable rates to premenopausal women (**Figure 3B)**. Even though hot flashes were the most reported symptom in the perimenopausal and menopausal cohorts, more menopausal users reported fatigue (75.0%) than hot flashes (73.1%) followed by reports of anxiety (58.7%), joint pain (56.1), and brain fog (56.1%).

Total symptom counts by user exhibited statistical differences between premenopausal, peri, and menopausal women. Aside from symptoms known to explicitly relate to either menopause or the menstrual cycle (i.e., vasomotor symptoms, ovulation and ovulation pain, cramps, spotting), several symptoms differed.

Premenopausal and perimenopausal women logged the largest differences in fatigue, bloating, headaches, diarrhea, and mood swings (p<0.01, chi-sq>48.7) compared to menopausal women. Menopausal women reported greatest differences in elevated rates of painful sex, insomnia, vaginal dryness, memory lapse, low sex drive, and uti (p<0.01, chi-sq>26). No differences were observed in anxiety or vertigo.

### Hierarchical Clustering of Symptom Covariance

#### Premenopausal

Premenopausal symptoms fell into mood/cognitive and digestive groups, with most other symptoms minimally covarying (**Supplemental Figure 3, top**). Brain fog and memory lapse were grouped more closely among regular cyclers, as were digestive and menstrual cycle-associated symptoms (e.g., breast pain, cramps, insomnia, bloating, constipation, ovulation pain) **(See Supplemental Figure 4)**. By contrast, the grouping of fatigue and a variety of mood and cognitive symptoms appeared more closely clustered in irregular cyclers **(See Supplemental Figure 4).**

#### Perimenopausal

Perimenopausal women exhibited 3 large symptom branches (**Supplemental Figure 3, Middle)**. The highest covarying symptom group included fatigue and mood/cognitive problems. Hot flashes were unrelated to any other symptoms.

#### Menopausal

Menopausal symptoms exhibited different structure from premenopausal or perimenopausal women **(Supplemental Figure 3)**, and further differed by type of menopause (natural vs. medical/surgical) (**Supplemental Figure 5**). In the menopause cohort as a whole, most data fell into a large, moderately correlated cluster including integumentary and mood/cognitive problems.

Mood/cognitive and integumentary problems were even more related in medical/surgically entered menopause **(Supplemental Figure 5).** Notably, hot flashes and night sweats were unrelated to any other symptoms within these hierarchies.

### K-Means Clustering of Symptom Covariance PCA

#### Premenopausal

Fifteen principal components (PCs) were needed to capture 87% of the variance in the premenopausal dataset. The first PC captured 23% of the variance, and the second PC a remaining 11%. PC 3 accounted for 9%, and PC4 7%. All remaining PCs were ≤5%. Relatively few symptoms contributed to the top PCs: spotting in PC1, fatigue, headaches, anxiety, bloating, cramps, and breast pain in PC2; breast pain and cramps PC3, headaches in PC4 (**See Supplemental Figure 2**). All observed premenopausal clusters shared a baseline phenotype resembling premenstrual syndrome. 81% of users fell into a cluster reporting fatigue, bloating, cramps, anxiety, and headache. A remaining 13% were differentiated by bloating. The remaining 6% exhibited a variety of integumentary complaints alongside spotting and additional digestive symptoms. The only observed difference in top symptoms for regular cyclers was in the presence of brain fog rather than headache in the most common cluster (76% of regular cyclers).

#### Perimenopausal

Fifteen (15) PCs were needed to capture 90% of the variance: 55% for PC1, 11% for PC2, and 5% for PC3 (**See Supplemental Figure 2**). Remaining PCs captured ≤ 3% of the variance (data not shown). PC 1 was exclusively determined by hot flashes, PC 2 was a mixture of many symptoms, PC 3 largely driven by nights sweats and, notably, the next PC driven by residual menstrual symptoms spotting and cramps. Top symptoms grouping each main segment are as follows: 81% of these women were placed in a segment characterized by VMS alongside fatigue, cramps, and bloating. 12% were characterized by their lack of night sweats, spotting, and headaches. The remaining exhibited additional digestive symptoms or muscular pain.

#### Menopausal

In menopausal women, 90% of the variance was captured in the first 3 PCs, the vast majority by PC1: 86% for PC1, 3% for PC2, and 3% for PC3 (**See Supplemental Figure 2**). Remaining PCs captured ≤ 3% of the variance (data not shown). PC 1 was almost exclusively determined by hot flashes, whereas PC2 was a mixture of fatigue, night sweats, mood/cognitive, and integumentary problems. PC3 was dominated by night sweats. The large majority (91%) of menopausal women were clustered by hot flashes and, similar to premenopausal women, fatigue, anxiety, bloating, and joint pain.

An additional 6% included night sweats. The remaining reported the above symptoms alongside insomnia, chills, and irregular heartbeat.

### Symptom Networks

Symptom networks varied greatly by hormonal stage of life. Premenopausal symptoms were linked more sparsely than perimenopausal or menopausal symptoms, and into 6 groups of 3 or more symptoms, with all remaining symptoms singletons or pairs. Groups were comprised of 1) cognitive/mood, 2) integumentary, 3) flu-like/digestive, 4) nervous/muscular pain, 5) sexual symptoms, and 6) menstrual cycle-associated. The nodes with highest degree centrality were brain fog, mood swings, dry skin, cramps, and nausea. For symptom networks by type of cycler, see (**Supplemental Figure 6**).

Perimenopausal symptoms also clustered into 6 similar groups of 3 or more symptoms, with remaining symptoms in dyads or alone (**Figure 4**). 1) cognitive/mood, 2) integumentary, 3) dizziness/vertigo/irregular heartbeat, 4) sexual symptoms, and two groups included menstrual cycle-associated symptoms (ovulation vs. premenstrual). Notably, hot flashes and night sweats clustered together but not with any other symptoms. Mood swings, constipation, hair loss, vaginal dryness, and depression exhibited the greatest degree centrality.

**Figure 4.**
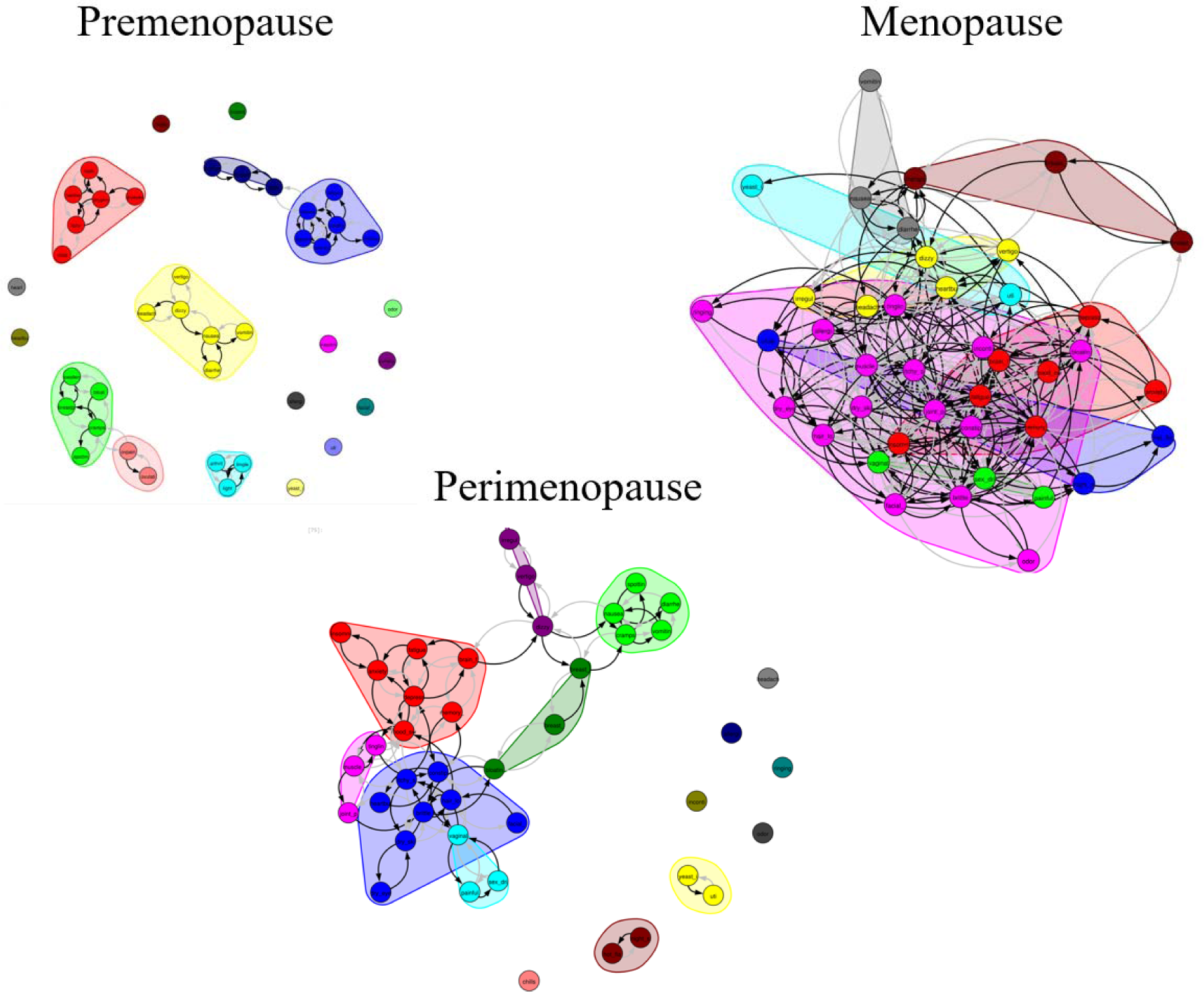
Premenopausal, perimenopausal, and menopausal symptom network analysis. Clusters are coded separately, and letters stand for each abbreviated symptom. For full key see: **Supplemental Table 1**.

Menopausal symptoms displayed a denser network than premenopausal symptoms, with similarly structured clusters to those found in HCA and K-Means clustering and some overlap with the premenopausal symptom network. Menopausal women maintained 1) cognitive/mood, 2) digestive and integumentary 3) dizziness/vertigo/irregular heartbeat similar to perimenopausal women, 4) sexual, 5) flu-like, and 6) hot flashes, night sweats and chills. Joint pain, fatigue, itchy and dry skin, and memory lapse exhibited highest degree centrality. For symptom networks by type of menopause, see (**Supplemental Figure 7**).

All symptom summaries across methods are described in (**Table 2**).

**Table 2.**
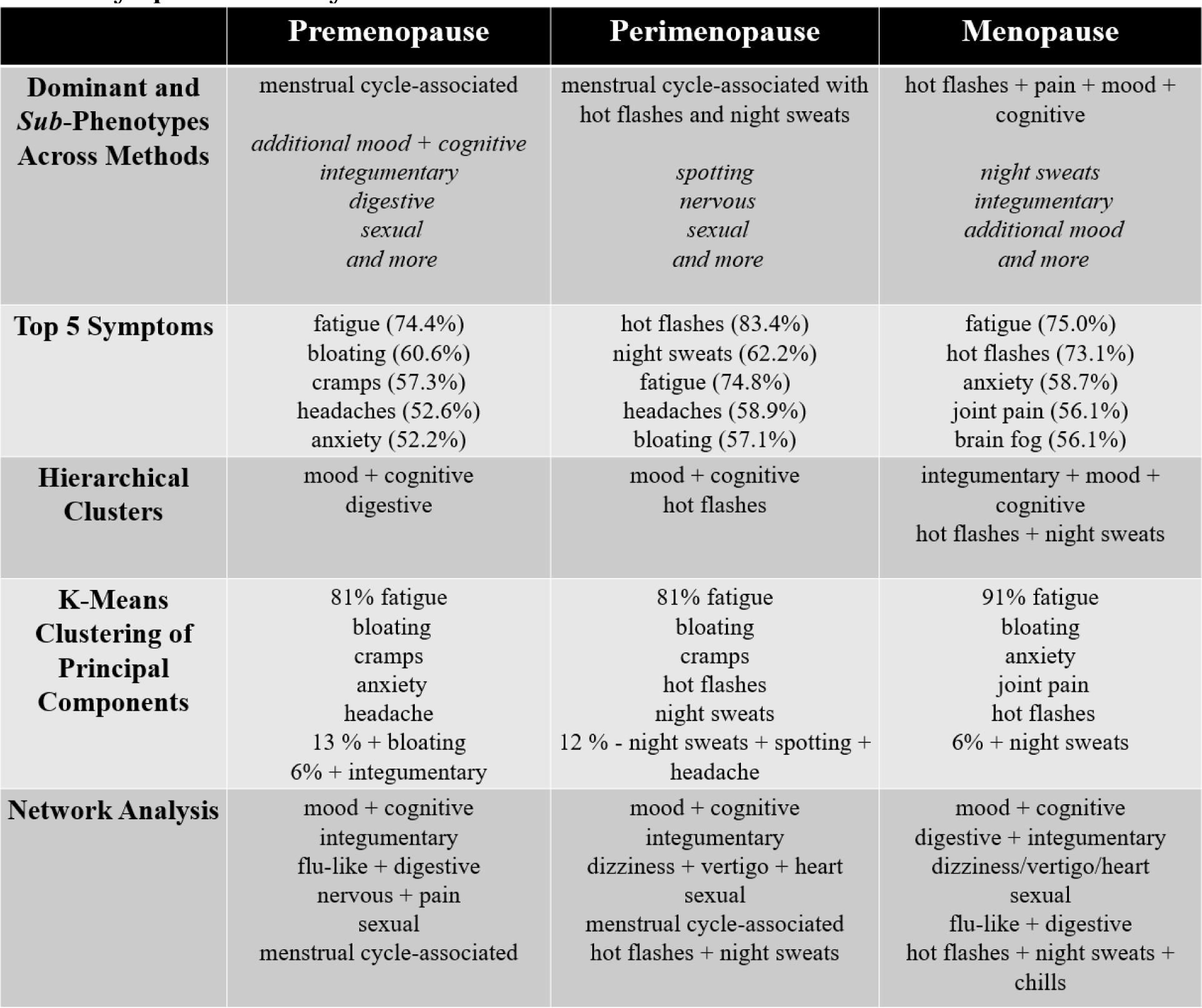
Symptom Summary Summary of symptom clusters across methods. Blue indicates shared symptom clusters or rankings across life stages, whereas red indicates differentiating symptoms and clusters.

## Discussion

Clustering of self-collected symptom data enabled construction of a detailed map of women’s experiences prior to, during, and following the transition to menopause. Each clustering method identified symptom groups resembling ovulation or PMS^36^ in premenopausal women, and classic VMS in menopausal women, whereas perimenopausal women exhibited a mixture of both symptom types. These were accompanied by a suite of symptoms that was shared across women regardless of hormonal status consisting of fatigue, bloating, joint pain, and anxiety. The symptoms above characterized ∼90% of each life stage cohort, with 1-2 phenotypes representing the remainder. Cycle and menopause type also impacted symptom relatedness, with regular cycles displaying greater symptom covariance. Those undergoing medical/surgical menopause, as expected, exhibited more diverse symptom clusters compared to those who experienced natural menopause. Finally, smaller clusters in each life stage appeared to relate to specific conditions, including integumentary and digestive complaints, allergies, mood/cognitive disturbance, sexual dysfunction, urinary tract infection, and yeast infection. It is remarkable that bloating, fatigue, and anxiety co-vary and cluster similarly and occur at nearly the same rate in premenopausal and menopausal women, despite typically being discussed as premenstrual symptoms. Interestingly, although these symptoms appear to persist from premenopausal to menopausal status; their frequency and tendency to cluster was greater in premenopausal women. Although new symptoms emerge with menopause, at least some recede.

In addition, the relation of common symptoms to one another changed by reproductive life stage. For instance, brain fog was strongly correlated and clustered with fatigue, headache, and other symptoms in menopausal women, but was relatively uncommon and unrelated to other symptoms in premenopausal women. Additionally, anxiety and depression exhibited higher covariance after menopause. Just as it is relevant what symptoms covary, it is just as interesting what prominent symptoms do not further predict an individual’s experience. Surprisingly hot flashes, occurring in about 80% of perimenopausal women and ¾ of the menopausal cohort, did not predict additional symptoms beyond night sweats. Similarly, reports linked to specific phases of the menstrual cycle: breast pain and swelling, spotting, cramps, headaches, ovulation pain – did not predict further symptom logs in any method.

Together, this cohort shared a common phenotype of fatigue, headache, anxiety, and brain fog that partially eased with age. Premenopausal women exhibited additional symptoms that are frequently attributable to the menstrual cycle (e.g., cramps, breast pain, spotting), whereas perimenopausal women exhibited these symptoms and VMS, and menopausal women reported VMS. Premenopausal subgroups were characterized by spotting in irregular cyclers and additional pain, skin, and mood symptoms, whereas menopausal subgroups were characterized by either night sweats or a subset of skin, additional mood/cognitive, or pain symptoms. Classic symptoms of VMS and PMS were minimally related to the symptoms a user logged.

Several previous studies have proposed symptom clusters in menopausal women, largely relying on in-clinic data collection during doctor’s appointments. These studies do not present a consistent picture of pre- or post-menopausal symptomology. Some studies identify clusters exclusively based on symptom severity (e.g., mildly, moderately, or severely symptomatic menopause) rather than symptom type^29^. Only a few studies directly compare premenopausal and menopausal cohorts. Although our study does not follow the same women across the menopausal transition, Harlow et al., 2017^10^ observed that the majority of women in the SWAN dataset stay in the same symptom class across menopause, consistent with our observation of a shared symptom suite across cohorts. Similar to our findings, Seib et al., 2017^24^ also identified VMS symptoms as a cluster separable from other symptoms. The causes for an individual’s symptom profile are unknown, but women with worst symptoms are more likely to have experienced financial stress, be white, be obese or have metabolic syndrome, and smoke^10^. Chinese and Japanese women also report fewer vasomotor symptoms, but still report fatigue, diffuse pain, digestive, sleep, respiratory, and psychological symptoms^27^. Our results may help reconcile these disparate findings, as we observed some differences in symptom cluster based on the method of analysis. Moreover, we identified a common symptom profile to ∼ 80-90% of women that existed in addition to each group’s traditional symptoms (PMS and VMS, or their mixture in pre-, peri-, and menopause, respectively).

The high frequency and relatedness of reported fatigue, headache, anxiety, and brain fog across both populations was unexpected. These symptoms are not typically associated with both younger and older age groups and may reflect both the self-selection of women who seek out a symptom tracking app, and the unique appeal of self-collected health data during the COVID-19 pandemic. However, symptoms of inflammation and cognitive/physical burnout have increased greatly in the past two decades, and this increase began well prior to the COVID-19 pandemic. The phenotype is attributed to many causes, including metabolic disruption caused by sweetened and processed food, to inter-related reproductive, metabolic, and sleep disruption, and to a high-stress American working culture. Whatever the specific contributors to this consistent symptom suite observed here, these symptoms represent a clinically relevant baseline of suboptimal health that may outweigh the burden of female symptoms that usually capture clinical focus and treatment. Strategies that prevent and treat fatigue, inflammation, and cognitive overwhelm in women of all ages would alleviate the majority of symptom burden for the majority of women, regardless of menopausal status.

### Self-Collected Data and Analytical Methods

This analysis relied on self-collected, rather than clinician-collected data, providing several advantages and limitations in terms of accuracy, detail, and timeliness. In contrast to in-clinic data collection, individuals in the Meno Life data set have the opportunity to collect repeat data over time. Most users interact with the app for ∼1 week, providing a representative view of their everyday life during this time as opposed to a recollected snapshot over many years. Self-collected data close to the time symptoms are experienced is likely to be more accurate than in-clinic recall once a year. Self-collected data may be particularly more accurate for symptoms which recur multiple times a day (e.g., hot flashes, chills), and which would otherwise be difficult to count and evaluate separately^37,38^. Finally, self-collected data using a mobile app on a personal smartphone may alleviate hesitation to report more personal symptoms (indeed, some previous studies omit questions about urogenital or sexual symptoms altogether in their survey^27^). This data is not without its downsides: without full medical histories, or clinician determination of menopausal status, we lack important background about participants and may classify some women incorrectly. Together, self-collected data typically provides more accurate, quantitative, and detailed information about symptoms. Even if some women may incorrectly report menopausal status, they are unlikely to consistently incorrectly report their personal experiences.

Beyond the method of data collection, clustering methods can generate different data groupings. For example, K-Means identifies clusters based on moving the location of a pre-specified number of centroids, whereas HAC and network analysis rely on the connectivity among data points and start by assuming that each symptom is its own cluster. Moreover, using K-Means to cluster principal components rather than all symptom data neglects symptoms that minimally contribute to variance in the dataset. Finally, network analysis uses binary input data, meaning that individuals who report more of the same symptoms do not exert a larger effect than individuals who report fewer of the same symptoms^39^.

We compared “top down” and “bottom up” clustering methods and identified consistent phenotypes common to all methods. We propose that the shared symptom groups may represent a broader “ground truth” of symptom relations in modern-day American women. Historical use of infrequently collected, recalled data, typical use of only one clustering method, use of data from 15+years ago, and use of data from very different populations may contribute to discrepancies in the determination of menopausal symptom clusters.

### Caveats and Limitations

Self-collected data from a symptom-tracking app has several limitations, including variable completeness and accuracy of symptom-logs, selection bias, unequal group size, and lack of medical records. First, although we omitted individuals with the greatest number of symptom logs (>300 symptoms), as well as individuals who reported too few symptoms to assess the covariance between 2 symptoms (<10), individuals who reported more symptoms have a stronger impact on symptom covariance than those who reported fewer symptoms. This does not impact network analysis but does impact hierarchical clustering and the PCA shown here. Second, symptom tracking apps are also most likely to attract individuals who have symptoms to report, meaning that this analysis may overestimate the prevalence of these symptoms in the general population. Even within the symptom-tracking population, those with higher symptom burdens may be more motivated to understand the patterning or lifestyle factors contributing to their symptoms, and therefore may log more regularly than less symptomatic users. Further, users may report the symptoms that they find interesting to track, and fail to report symptoms which they are not interested in. In each of these scenarios, the symptom frequencies reported here are potentially an over-representation of “compelling” symptom burden in the general female population. Finally, this study lacks detailed demographic information, including ethnicity, socioeconomic status, pre-existing conditions and medications; all of which contribute to symptom burden^10,11,24,27^. Lack of clinical oversight means that menopausal status was not verified by a clinician, some individuals may have mis-reported status during onboarding (e.g., reporting a menstrual period when menses were absent). Although users who reported conflicting information were omitted, errors are still possible. Separating pre from perimenopausal users was also not possible using self-reports of period data alone, as we did not assess ovulation regularity or hormonal patterns.

## Conclusions

Together, clustering of self-collected symptom data in American women from pre to post menopause revealed a common phenotype of fatigue, headache, brain fog, and anxiety; alongside cycle-associated symptoms in pre- and perimenopause, and VMS in peri- and menopause. These were accompanied by diverse additional digestive, integumentary, mood, nervous, and sexual symptoms in a subset of women. Symptom burden in women’s health extends far beyond symptoms associated with reproductive status, and such self-collected data entry tools create a future opportunity to evaluate impact and effectiveness of both hormonal, dietary, and targeted interventions on symptom clusters.

## Methods

### Data Collection and Inclusion Criteria

All procedures have been approved by Western Institutional Review Board-Copernicus Group (registration number, OHRP and FDA, IRB0000053; parent organization number, IORG0000432) *Study Number: 1284093*. Anonymized data were drawn from the MenoLife mobile app created by MenoLabs (https://app.menolabs.com) and collected between Fall 2021 and Spring 2023. Users completed an onboarding questionnaire that included whether menstruation occurred in the last twelve months, description of menstrual periods, birth control status, how the user entered menopause (if they noted absence of menstrual periods or > 12 months since last menstruation) and, finally, selection of most common symptoms. Following onboarding, women used the app at will to enter symptom logs from a list of 45 available symptoms.

Onboarding data was used to estimate user status as premenopausal or menopausal. These groups were separated for further analysis. Briefly, if users indicated that they had not entered menopause, and further specified that they had had a menstrual period within 12 months, and did not record vasomotor symptoms (i.e., hot flashes or night sweats), they were classified as premenopausal. Users were further grouped by whether they reported regular or irregular cycles. If users stated that they had entered menopause, and further confirmed that they had not had a period in more than 12 months, they were classified as menopausal. Menopausal users were further grouped by whether they reported entering menopause naturally, or via medical/surgical interventions. Individuals reporting chemotherapy (n=6) were not included. Perimenopause does not have a strict clinical definition using symptoms alone. Here we chose to estimate the perimenopausal population as users that indicated they had not entered menopause, who reported irregular cycles within the past year, and who experienced vasomotor symptoms. Users that selected conflicting answers were omitted from further analysis (e.g., self-identified as menopausal but selected that their periods were regular). As individual onboarding questions could be skipped, and multiple answers could be selected in some cases to each question, users of indeterminate status were also omitted from further analysis.

Finally, as many users were minimally interactive with the app, logging only a few symptoms, we opted to include only users who had logged at least 10 symptoms for further analysis of relationship among symptoms. To minimize the impact of “super-users” on symptom covariance, we opted to omit individuals who had logged >300 symptoms.

### Symptom Categories

Menstrual cycle associated symptoms were here defined as ovulation and ovulation pain, menstrual cramps, breast pain and swelling. Although fatigue and mood changes are commonly considered premenstrual symptoms, we considered these separately as they were characteristic of both pre and post-menopausal women. Vasomotor symptoms were defined as hot flashes and night sweats.

Although chills may be defined as VMS, they can also result from illness/infection, and so were considered separately. Integumentary symptoms refer to symptoms affecting hair, skin, and nails.

### Data Analysis and Statistics

Data were securely organized in Amazon Web Services (AWS) S3 and queried through AWS Athena. Custom Python and R code was written for all the analysis methods. Ranksum tests (non-parametric ANOVA) were used to avoid assumptions of normality in comparisons of symptom count by individual across all 45 measured symptoms. Prior to computing the covariance matrix, symptom counts were standardized across symptoms rather than across users, meaning that users who experienced more symptoms contributed more strongly to hierarchical agglomerative clustering (HAC) (Python: clustermap() from *seaborn*, linkages generated using linkage() from *scipy.cluster.hierarchy*).

#### Hierarchical Clustering Analysis

HCA combines independent forests of clusters that are not part of an existing hierarchy by using a distance metric to grow clusters. It starts by treating each symptom as a single node “forest”, maintaining a distance matrix between all clusters. This distance matrix is updated at each bottom-up iteration, with the algorithm converging at the formation of a single cluster. The distance metric used is min(dist(u[i], v[j])) where u, v are 2 clusters, and i, j represents each element within that cluster. This is repeated for all pairs of clusters up the hierarchical chain. We use Euclidean distance for dist() function. Dendrograms representing the hierarchical structure of symptom data were generated using the unweighted pair group method using arithmetic mean (UPGMA) applied to the covariance matrix of normalized symptom counts, and silhouette score was used to determine clusters reported here.

#### Principal Components Analysis

We used PCA (Python: PCA() from *sklearn.decomposition*) to reduce the dimensionality of the symptom dataset prior to K-Means clustering. K-Means clusters were generated using the principal components needed to capture 90% of the variance in each group’s symptom data (9 principal components (PCs) for premenopausal data, 3 PCs for menopausal data and 13 for perimenopause), and elbow of the sum of squared errors (SSE) and silhouette score were used to determine optimal cluster number.

#### K-Means Clustering

K-Means clustering of principal components generated from normalized symptom count was used to evaluate potential consensus with hierarchical clustering. Recall that HCA is a bottom-up approach in which each symptom-pair began as its own cluster, and clusters are iteratively merged or split until all points have been accounted for. By contrast, K-Means partitions data into a set number of clusters and aims to place data points into the group with the nearest centroid. We aimed to compare results generated under these methods to identify what symptom clusters were identified in both, as well as any small but notable groupings identified by HAC (e.g., the grouping of mood and cognitive symptoms).

#### Network Analysis

Network analysis was performed in R using the package IsingFit^40,41^. The network estimation procedure used, called “eLasso” and based on the Ising model, pairs regularized logistic regression with model selection based on the Extended Bayesian Information Criterion (EBIC), a measure of fit that identifies variable relationships of interest. The resulting network consists of a symmetric (undirected) weight adjacency matrix. Each value above (below) the diagonal represents an edge (relationship) between a variable in a given row to the variable in that column.

Input data to Isingfit were one hot encodings of the symptom matrices from each group. The presence of a symptom in any count in each individual was converted to a 1, and absence of a symptom remained a zero. Symptoms were set to null for which values were either (a) all blank, (b) rare enough that the Isingfit reported error due to lack of co-variance. These were nipple discharge in all groups; hot flashes and night sweats for premenopausal women; and ovulation, ovulation pain, spotting, and vomiting for menopausal women.

Networks were then exported and visualized in Python using the package iGraph. The Walktrap algorithm (Python: walktrap() from *iGraph*) was used to identify relevant communities within the network^42^.

Negative correlations could not be used as inputs, were rare in the networks estimated by Isingfit, and were removed. Walk trap was tested with 3-10 steps, and the number of steps minimally impacted the estimated communities. Graphs displayed use 4 steps. Plots shown depict detected communities as shaded and nodes belonging to those communities in the same color. For a list of symptom names and abbreviations displayed on the graphs see Supplemental Table 2.

Node degree, betweenness, closeness, and strength were calculated using iGraph (Python: degree(), betweenness(), closeness(), and strength () from *iGraph*) and used to identify the most important nodes in the network. Degree is the number of edges connected to a given node, betweenness is the extent to which one node lies along the shortest path between other nodes, closeness is a measure of the average path length between one node and the others in the network, and strength is the sum of weights attached to ties belonging to a given node.

## Author Contributions

Study conception and data collection: SA, JK. Hypothesis generation. SA, AG, JK. Data analysis: SA and AG. Writing: AG, JK, SA. All authors read and approved the final manuscript.

## Data and Code Availability

Python and R code used for analysis is openly available as Jupyter notebooks at the following Github repository - https://github.com/nextgensh/amyris-analysis.git. Raw data is available on request from the authors.

## Supplemental Figures & Tables

**Supplemental Figure 1.**
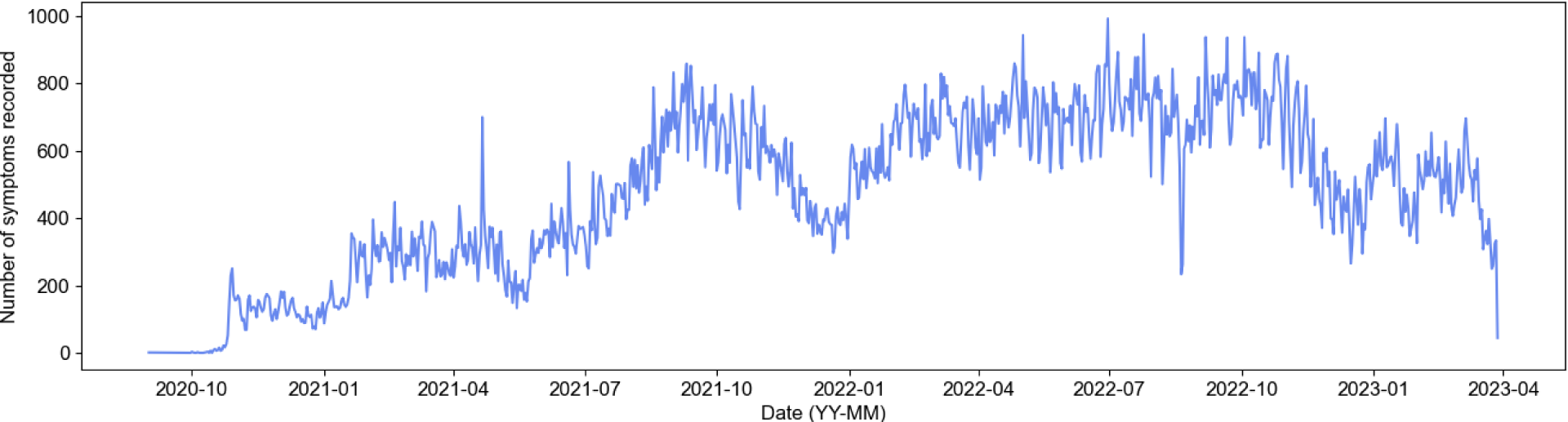
Number of symptoms collected through the app over the years. We used symptoms from Fall 2021 to Spring 2023 in our analysis. We see a seasonal trend in symptom recording, with compliance dropping

**Supplemental Figure 2.**
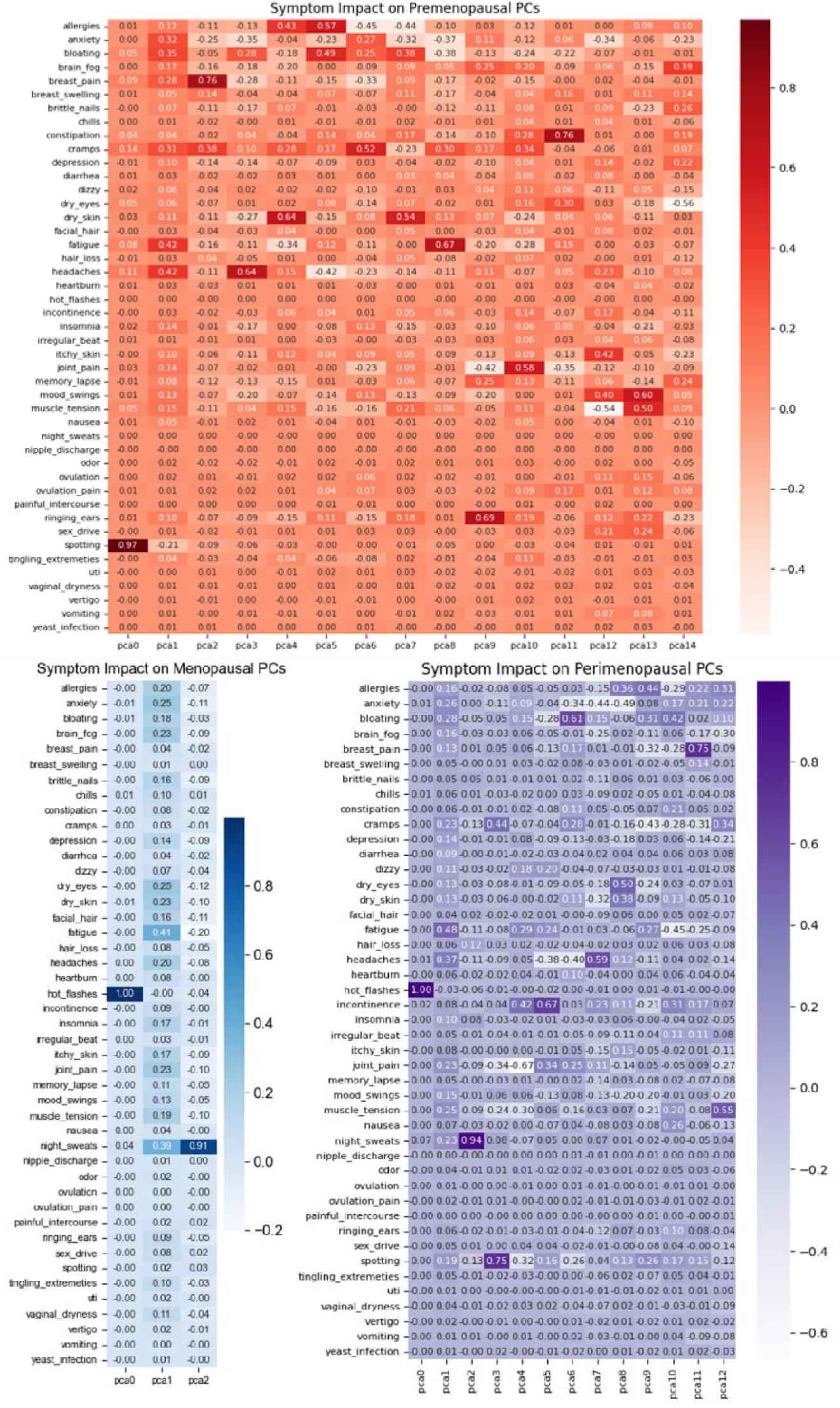
Impact of each symptom on principal components in pre-(red), peri-(purple), and menopausal (blue) women.

**Supplemental Figure 3.**
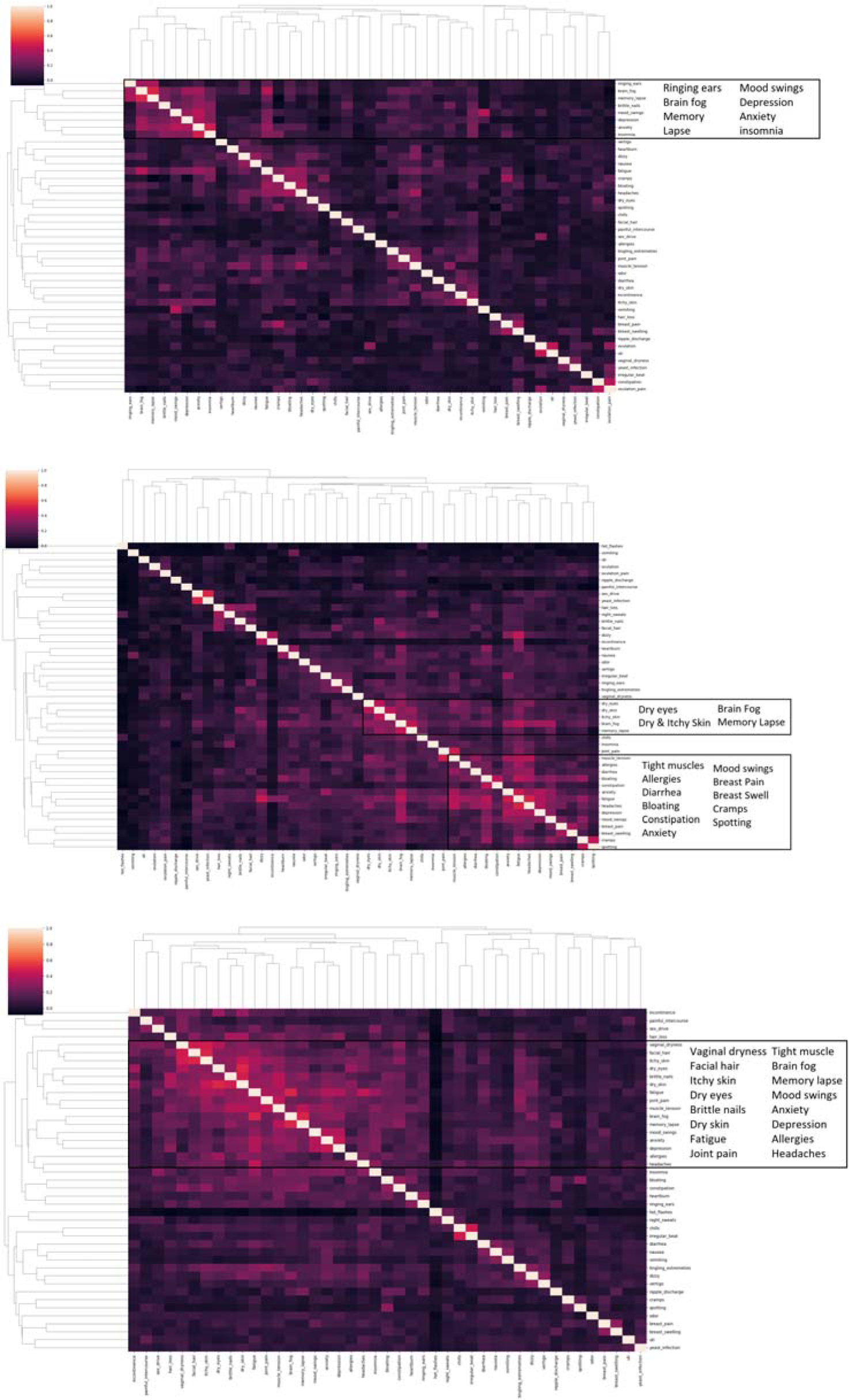
Hierarchical cluster maps in premenopausal (top), perimenopausal (middle), and menopausal (bottom) participants.

**Supplemental Figure 4.**
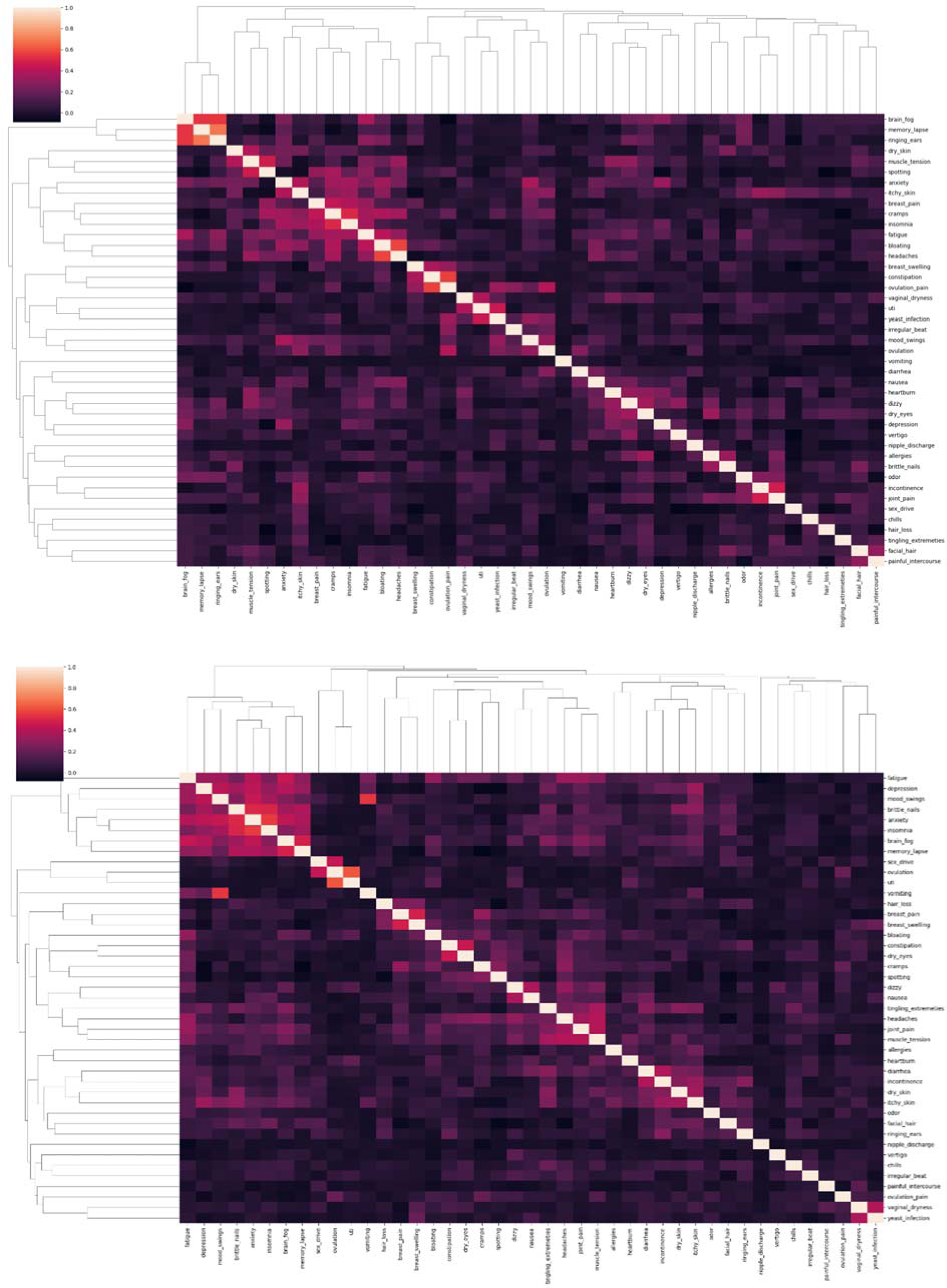
Hierarchical cluster maps in regular cyclers (top) and irregular cyclers (bottom).

**Supplemental Figure 5.**
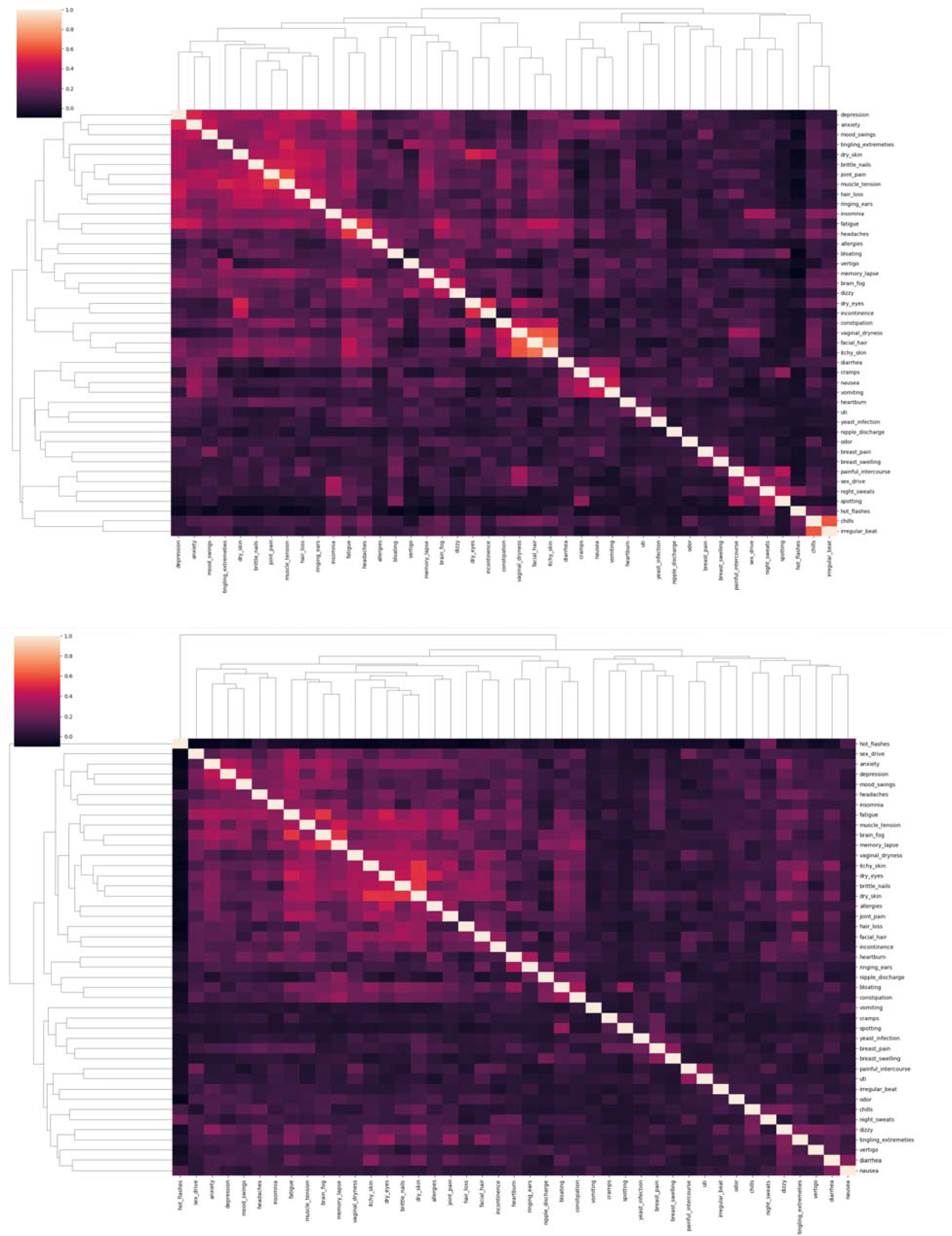
Hierarchical cluster maps in medical/surgical menopause (top) and natural menopause (bottom).

**Supplemental Figure 6.**
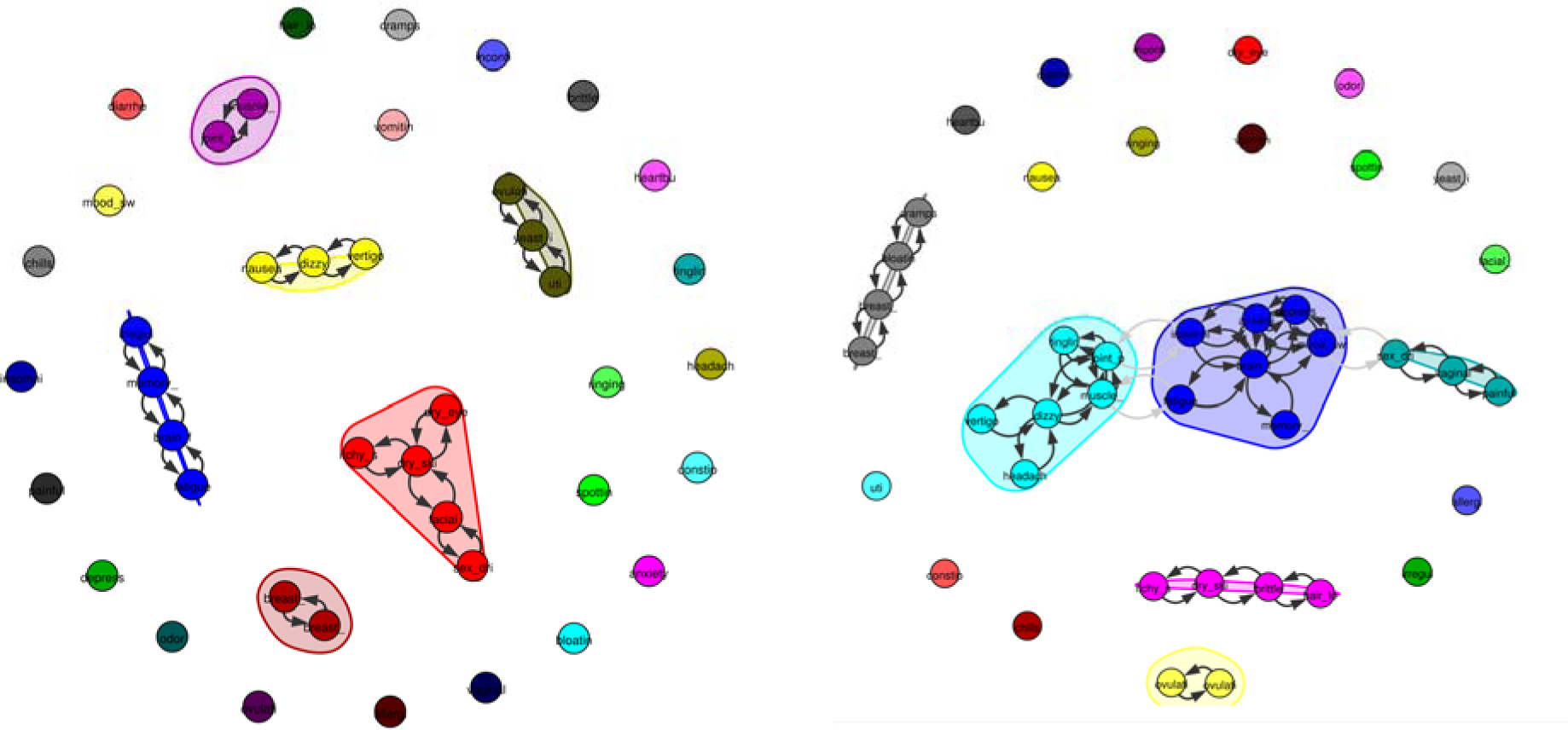
Regular (left) and irregular (right) cyclers’ symptom networks. Symptom groups were more related in irregular cyclers, with pain/nervous, mood/cognitive, and sexual clusters linked.

**Supplemental Figure 7.**
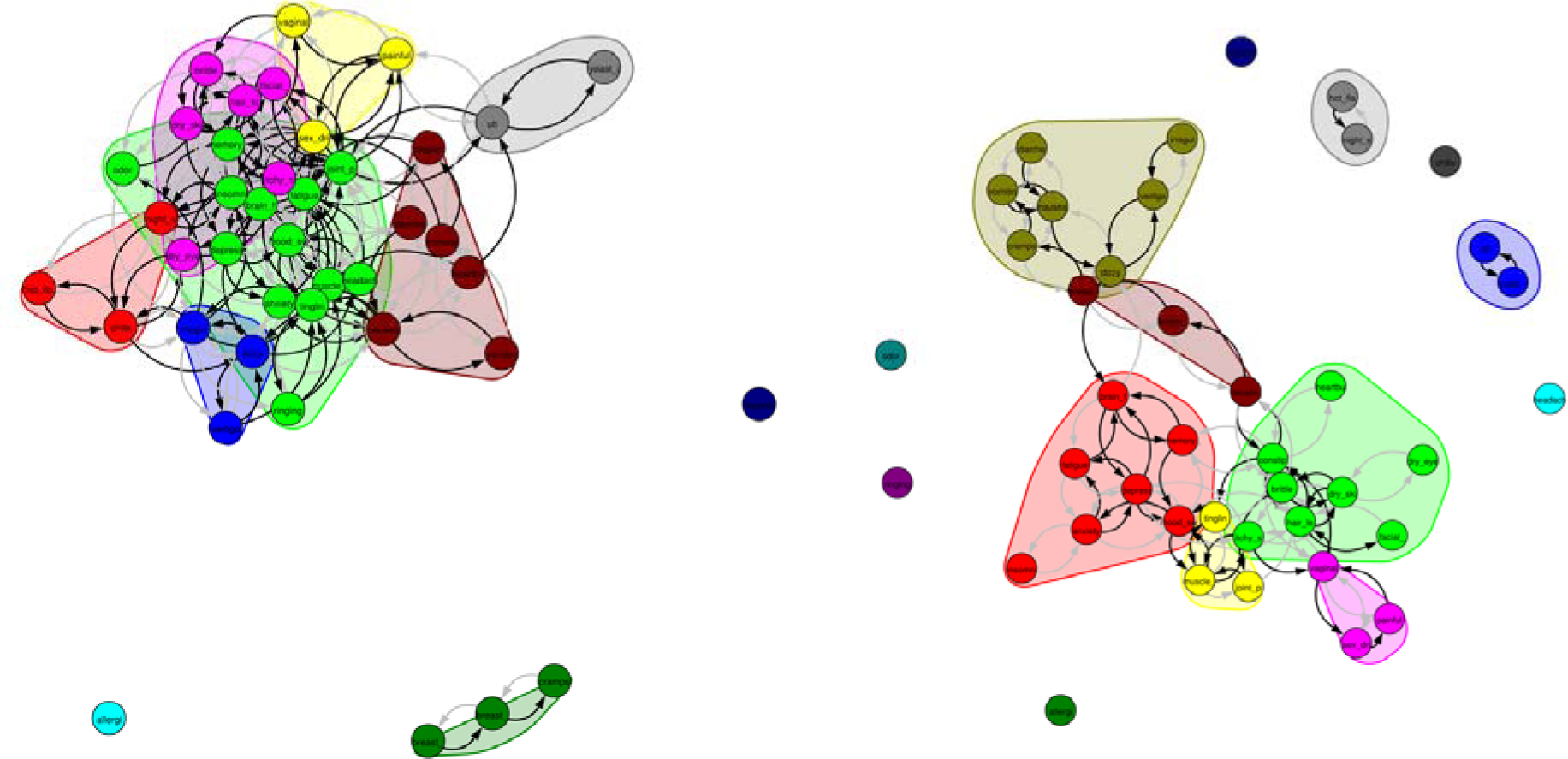
Natural menopausal users (left) exhibited a more connected symptom network than did medical/surgical. Notably, hot flashes and night sweats were not connected to any additional symptoms in medical/surgical hysterectomy but were connected to a variety of mood and nervous symptoms in natural menopause.

**Supplemental Table 1.**
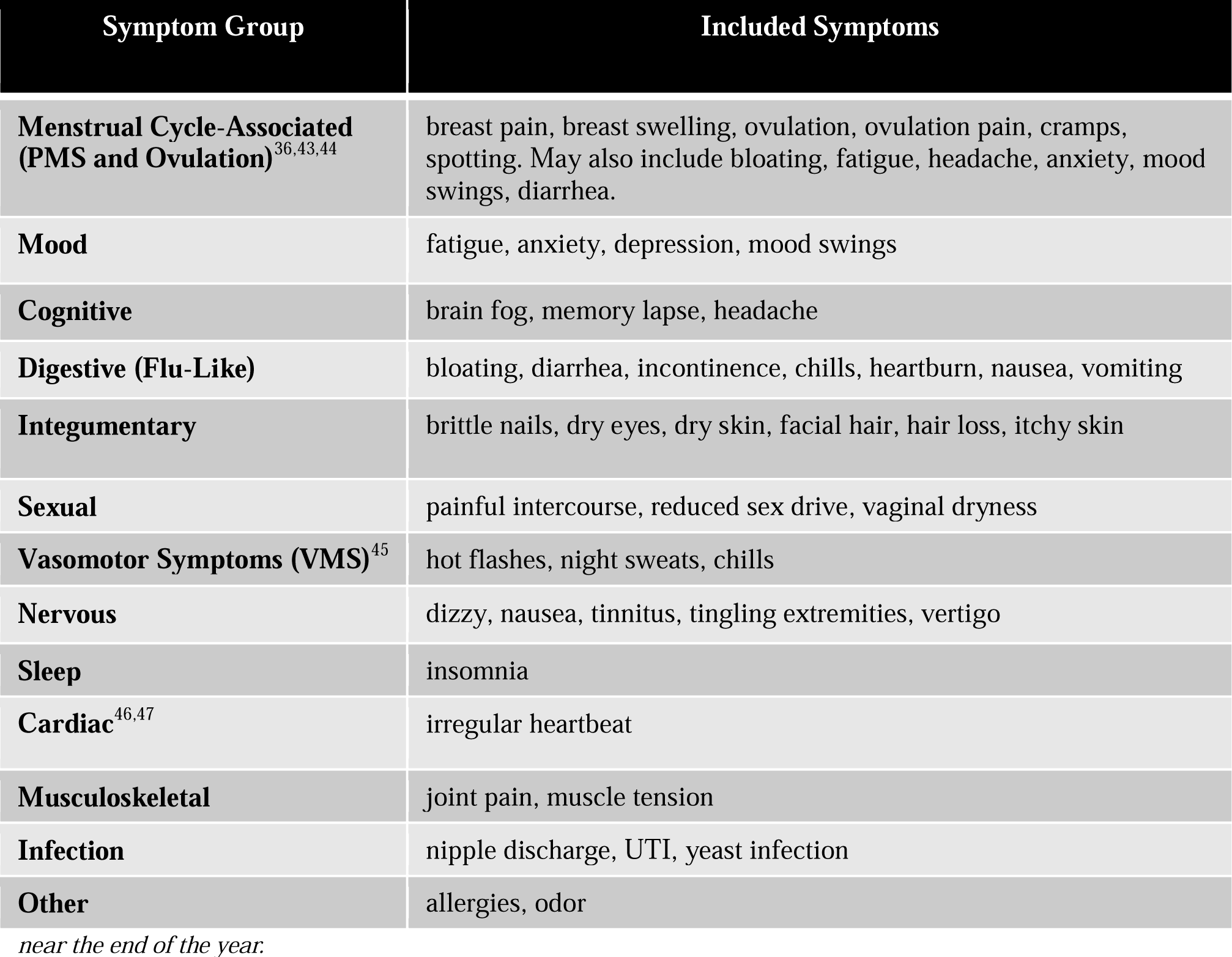
Symptom Group Descriptions.

## References

1. Singh, A., Kaur, S. & Walia, I. A historical perspective on menopause and menopausal age. Bull. Indian Inst. Hist. Med. Hyderabad 32, 121–135 (2002).

2. Richardson, J. T. The premenstrual syndrome: a brief history. Soc. Sci. Med. 1982 41, 761–767 (1995).

3. CDC Data, F. Health, United States 2019. CDC https://www.cdc.gov/nchs/hus/data-finder.htm?year=2019&table=Table%20026 (2019).

4. 4. Obesity is a Common, Serious, and Costly Disease. *Centers for Disease Control and Prevention* https://www.cdc.gov/obesity/data/adult.html (2022).

5. 5. NASH Definition & Prevalence - American Liver Foundation. https://liverfoundation.org/liver-diseases/fatty-liver-disease/nonalcoholic-steatohepatitis-nash/nash-definition-prevalence/.

6. Moore, J. X. Metabolic Syndrome Prevalence by Race/Ethnicity and Sex in the United States, National Health and Nutrition Examination Survey, 1988–2012. Prev. Chronic. Dis. 14, (2017).

7. Grelle, K. et al. The Generation Gap Revisited: Generational Differences in Mental Health, Maladaptive Coping Behaviors, and Pandemic-Related Concerns During the Initial COVID-19 Pandemic. J. Adult Dev. 1–12 (2023) doi:10.1007/s10804-023-09442-x.

8. Vidal-Cevallos, P., Mijangos-Trejo, A., Uribe, M. & Tapia, N. C. The Interlink Between Metabolic-Associated Fatty Liver Disease and Polycystic Ovary Syndrome. Endocrinol. Metab. Clin. North Am. 52, 533–545 (2023).

9. Phumsatitpong, C., Wagenmaker, E. R. & Moenter, S. M. Neuroendocrine interactions of the stress and reproductive axes. Front. Neuroendocrinol. 63, 100928 (2021).

10. Harlow, S. D. et al. It is not just menopause: symptom clustering in the Study of Women’s Health Across the Nation. Womens Midlife Health 3, 2 (2017).

11. Min, S. H. et al. Identification of high-risk symptom cluster burden group among midlife peri- menopausal and post-menopausal women with metabolic syndrome using latent class growth analysis. Womens Health 19, 17455057231160955 (2023).

12. Liu, H.-F., Meng, D.-F., Yu, P., De, J.-C. & Li, H.-Y. Obesity and risk of fracture in postmenopausal women: a meta-analysis of cohort studies. Ann. Med. 55, 2203515 (2023).

13. Martínez-Vázquez, S., Hernández-Martínez, A., Peinado-Molina, R. A. & Martínez-Galiano, J. M. Impact of overweight and obesity in postmenopausal women. Climacteric J. Int. Menopause Soc. 1–6 (2023) doi:10.1080/13697137.2023.2228692.

14. Grigsby, T. J., Howard, K., Howard, J. T. & Perrotte, J. COVID-19 Concerns, Perceived Stress, and Increased Alcohol Use Among Adult Women in the United States. Clin. Nurs. Res. 32, 84–93 (2023).

15. Zimmerman, M. E. et al. COVID-19 in the Community: Changes to Women’s Mental Health, Financial Security, and Physical Activity. AJPM Focus 2, 100095 (2023).

16. United States: life expectancy 1860-2020. *Statista* https://www.statista.com/statistics/1040079/life-expectancy-united-states-all-time/.

17. Kim, M. Y., Im, S.-W. & Park, H. M. The Demographic Changes of Menopausal and Geripausal Women in Korea. J. Bone Metab. 22, 23–28 (2015).

18. Belanger, H. G., Lee, C. & Winsberg, M. Symptom clustering of major depression in a national telehealth sample. J. Affect. Disord. 338, 129–134 (2023).

19. Meijs, C. et al. Identifying distinct clinical clusters in heart failure with mildly reduced ejection fraction. Int. J. Cardiol. 386, 83–90 (2023).

20. Niimi, N. et al. Which congestion presentation pattern on the physical findings is associated with future adverse events? A cluster analysis in the multicenter acute heart failure registry. Clin. Res. Cardiol. Off. J. Ger. Card. Soc. 112, 1108–1118 (2023).

21. Johansen, I. et al. Symptoms and symptom clusters in patients newly diagnosed with inflammatory bowel disease: results from the IBSEN III Study. BMC Gastroenterol. 23, 255 (2023).

22. 22. Pierson, E., Althoff, T. & Leskovec, J. Modeling Individual Cyclic Variation in Human Behavior. Preprint at 10.48550/arXiv.1712.05748 (2018).

23. Siegel, J. P., Myers, B. J. & Dineen, M. K. Premenstrual tension syndrome symptom clusters. Statistical evaluation of the subsyndromes. J. Reprod. Med. 32, 395–399 (1987).

24. Seib, C. et al. Menopausal symptom clusters and their correlates in women with and without a history of breast cancer: a pooled data analysis from the Women’s Wellness Research Program. Menopause 24, 624 (2017).

25. Gehlert, S., Chang, C.-H. & Hartlage, S. Symptom Patterns of Premenstrual Dysphoric Disorder as Defined in the Diagnostic and Statistical Manual of Mental Disorders-IV. J. Womens Health 8, 75–85 (1999).

26. Silva, C. M. L. da, Gigante, D., Carret, M. L. V. & Fassa, A. [Population study of premenstrual syndrome]. Rev. Saude Publica (2006).

27. Ho, S. C. et al. Menopausal symptoms and symptom clustering in Chinese women. Maturitas 33, 219–227 (1999).

28. Im, E.-O., Ko, Y. & Chee, W. Ethnic Differences in the Clusters of Menopausal Symptoms. Health Care Women Int. 35, 549–565 (2014).

29. Woods, N. F., Cray, L., Mitchell, E. S. & Herting, J. R. Endocrine biomarkers and symptom clusters during the menopausal transition and early postmenopause: observations from the Seattle Midlife Women’s Health Study. Menopause 21, 646 (2014).

30. 30. Ainsworth, A. J., et al. Global Menstrual Cycle Symptomatology as Reported by Users of a Menstrual Tracking Mobile Application. 2022.10.20.22280407 Preprint at 10.1101/2022.10.20.22280407 (2023).

31. Hantsoo, L. et al. Premenstrual symptoms across the lifespan in an international sample: data from a mobile application. Arch. Womens Ment. Health 25, 903–910 (2022).

32. SWAN Study Data Access - Women’s Health Across the Nation. *SWAN - Study of Women’s Health Across the Nation* https://www.swanstudy.org/swan-research/data-access/.

33. Im, E.-O., Lee, B., Chee, W., Brown, A. & Dormire, S. Menopausal Symptoms Among Four Major Ethnic Groups in the U.S. West. J. Nurs. Res. 32, 540–565 (2010).

34. Richard-Davis, G. & Wellons, M. Racial and Ethnic Differences in the Physiology and Clinical Symptoms of Menopause. Semin. Reprod. Med. 31, 380–386 (2013).

35. Grisso, J. A., Freeman, E. W., Maurin, E., Garcia-Espana, B. & Berlin, J. A. Racial Differences in Menopause Information and the Experience of Hot Flashes. J. Gen. Intern. Med. 14, 98–103 (1999).

36. Ryu, A. & Kim, T.-H. Premenstrual syndrome: A mini review. Maturitas 82, 436–440 (2015).

37. van de Belt, T. H., et al. Barriers to and Facilitators of Using a One Button Tracker and Web-Based Data Analytics Tool for Personal Science: Exploratory Study. JMIR Form. Res. 6, e32704 (2022).

38. Nahum-Shani, I. et al. Just-in-Time Adaptive Interventions (JITAIs) in Mobile Health: Key Components and Design Principles for Ongoing Health Behavior Support. Ann. Behav. Med. Publ. Soc. Behav. Med. 52, 446–462 (2017).

39. Jing, F. et al. Contemporaneous symptom networks and correlates during endocrine therapy among breast cancer patients: A network analysis. Front. Oncol. 13, 1081786 (2023).

40. Dalege, J., Borsboom, D., van Harreveld, F. & van der Maas, H. L. J. Network Analysis on Attitudes: A Brief Tutorial. Soc. Psychol. Personal. Sci. 8, 528–537 (2017).

41. Borkulo, C. van, Epskamp, S. & Robitzsch, with contributions from A. IsingFit: Fitting Ising Models Using the ELasso Method. (2016).

42. Pons, P. & Latapy, M. Computing communities in large networks using random walks.

43. Clayton, A. H. Symptoms related to the menstrual cycle: diagnosis, prevalence, and treatment. J. Psychiatr. Pract. 14, 13–21 (2008).

44. Brott, N. R. & Le, J. K. Mittelschmerz. in StatPearls (StatPearls Publishing, 2023).

45. Thurston, R. C. & Joffe, H. Vasomotor Symptoms and Menopause: Findings from the Study of Women’s Health Across the Nation. Obstet. Gynecol. Clin. North Am. 38, 489–501 (2011).

46. Enomoto, H. et al. Independent association of palpitation with vasomotor symptoms and anxiety in middle-aged women. Menopause N. Y. N 28, 741–747 (2021).

47. Carpenter, J. S. et al. Correlates of palpitations during menopause: A scoping review. Womens Health 18, 17455057221112267 (2022).

